# Changes in Eosinophil Counts and Glycemic Control in Patients with Diabetes

**DOI:** 10.64898/2026.10.15.26365690

**Authors:** Sujay Banerjee, Fritha Morrison, Maria Shubina, Alexander Turchin

## Abstract

Glycemic control is key to prevention of complications of diabetes. Although eosinophils have been implicated in metabolic regulation, most human studies have been cross-sectional and have yielded inconsistent findings.

We conducted a retrospective cohort study of adults with uncontrolled diabetes receiving primary care at Mass General Brigham. We evaluated the relationship between eosinophil count and HbA1c over one year, adjusting for demographic, socioeconomic, and clinical covariates.

Among 21,447 patients included in the study, eosinophil count increased by a mean of 2.6 cells/μL (SD 157.2 cells/μL) and HbA1c decreased by a mean of 0.82% (SD 2.09%) during follow-up. Increases in eosinophil count were associated with reductions in HbA1c (parameter estimate −0.366 per 1,000 cells/μL; 95% CI −0.526 to −0.206; P < 0.001), whereas baseline eosinophil count was not significantly associated with HbA1c changes. The association remained significant in a sensitivity analysis excluding systemic glucocorticoid users.

Increases in eosinophil count were associated with improved glycemic control in adults with diabetes, whereas baseline eosinophil count was not. These findings extend prior cross-sectional observations and suggest that changes in eosinophils may represent a biomarker of glycemic improvement. Further prospective studies are needed to determine whether this relationship reflects underlying immune-metabolic mechanisms or treatment-related changes.

## Introduction

Diabetes mellitus is a major cause of morbidity and mortality, and persistent hyperglycemia is strongly associated with microvascular and macrovascular complications (1, 2). HbA1c is the standard clinical measure of long-term glycemia and remains central to assessment of diabetes control in both research and routine care (1). Identifying biologic markers associated with longitudinal changes in HbA1c may improve understanding of heterogeneity in disease progression and treatment response among adults with diabetes (1, 2).

Eosinophils have traditionally been studied in allergic and parasitic disease, but they are increasingly recognized as tissue-resident immune cells with potential roles in metabolic regulation (3, 4). Experimental studies have suggested that eosinophils help modulate inflammation within adipose tissue and support insulin sensitivity and glucose homeostasis, although subsequent studies indicate that these effects depend on tissue context and cellular function rather than eosinophil abundance alone (4-6). Human observational findings have also been inconsistent. One large cross-sectional study reported that higher eosinophil counts were associated with lower prevalence of type 2 diabetes and less insulin resistance, whereas another population-based study found higher eosinophil counts to be associated with higher HbA1c, BMI, obesity, and metabolic syndrome, but not with fasting glucose, type 2 diabetes, or blood pressure (7, 8). These results suggest that circulating eosinophil counts may be related to metabolic health, but the direction and clinical significance of these relationships remain uncertain.

Despite this emerging evidence, little is known about the relationship between changes in eosinophil count over time and longitudinal glycemic control. Most prior human studies have been cross-sectional and have examined baseline eosinophil levels in relation to prevalent diabetes, insulin resistance, or other metabolic traits rather than subsequent change in HbA1c (7, 8). We therefore conducted a retrospective cohort study of adults with diabetes to evaluate the relationships of baseline eosinophil count and change in eosinophil count with change in HbA1c over time.

## Methods

### Study Design

We performed a retrospective cohort study to assess the relationship between changes in eosinophil counts and glycemic control in adults with diabetes. This study was approved by the Mass General Brigham institutional review board and the requirement for informed consent was waived.

### Study Cohort

We included in the study adults (age ≥ 18 years) with uncontrolled diabetes receiving primary care in the Mass General Brigham system between January 1, 2000 and June 30, 2022 who had sufficient longitudinal data for analysis. Specifically, patients were required to have at least one primary care note (to ensure that they had comprehensive documentation of their medical history in Mass General Brigham electronic medical records); a diagnosis of diabetes; a baseline HbA1c ≥7.0% (53 mmol/mol) (to be able to evaluate glycemic changes in both directions), a baseline and a follow-up HbA1c measurement; and a baseline and a follow-up eosinophil count. Patients entered the study on the first date (Index Date) when they fulfilled all eligibility criteria. Patients were excluded from the analysis if they had missing demographic information (biological sex or Social Deprivation Index / SDI (9)) or if their baseline eosinophil count was potentially indicative of a myeloproliferative process (>1,500 cells/μL).

### Study Measurements

Demographic characteristics, diagnoses, encounter data, insurance, laboratory data, and medication records were obtained from the electronic health record at Mass General Brigham. Diagnoses were identified using ICD codes. SDI was determined based on the patient’s zip code(9). Charlson Comorbidity Index (10) was calculated from diagnosis data available up to study entry.

An individual patient served as the unit of analysis. The primary outcome was the change in HbA1c from baseline to the end of the 1-year follow-up period after the Index Date. Baseline HbA1c was defined as the most recent HbA1c measurement within 3 years prior to or on the Index Date. Follow-up HbA1c was defined as the HbA1c measurement closest 1 year after the Index Date, among measurements obtained between 12 and 24 months after the Index Date.

Baseline eosinophil count was defined as the most recent measurement within 3 years before the Index Date; if no pre-Index Date measurement was available, the first measurement within 30 days after the Index Date was used. Follow-up eosinophil count was defined as the measurement closest to 1 year after the Index Date, among measurements obtained between 12 and 24 months after the Index Date.

Baseline systolic blood pressure and weight were defined as the most recent measurements within 3 years before or on the Index Date; if no pre-Index Date measurement was available, the first measurement within 30 days after the Index Date was used. Follow-up systolic blood pressure and weight were defined as the measurements closest to 1 year after the Index Date, among measurements obtained between 12 and 24 months after the Index Date. Change in systolic blood pressure and weight was defined as follow-up minus baseline.

### Statistical Analysis

Descriptive statistics were calculated using frequency counts for categorical and binary variables and summary statistics for continuous variables. ANOVA was used to compare change in HbA1c across quartiles of the change in eosinophil count. To conduct an adjusted analysis evaluating the relationship between the change in eosinophil counts and change in HbA1c over the follow-up period, we constructed multivariable linear regression models that were fit using HbA1c change from baseline to follow-up as the dependent variable and age, sex, race, preferred language, marital status, government insurance, SDI, veteran status, BMI, baseline HbA1c, Charlson Comorbidity Index, baseline eosinophil count, and change in eosinophil count from baseline to the end of the follow-up period as independent variables. We also conducted a subgroup analysis stratified by age (<65 vs. ≥65 years) for the primary HbA1c model.

In a sensitivity analysis, we excluded all patients who used systemic glucocorticoids either in the year prior to the Index Date or during the follow-up period and repeated the primary multivariable analysis in this restricted cohort. As secondary analyses, we evaluated the relationships between eosinophil count change and change in systolic blood pressure and weight. For each outcome, we compared mean change across quartiles of eosinophil count change using ANOVA and then fit multivariable linear regression models analogous to the primary HbA1c analysis, restricting each analysis to patients with available baseline and outcome measurements.

In all analyses missing BMI values were handled with 10-fold multiple imputation, where applicable. All analyses were performed using SAS 9.4 (Cary, NC).

## Results

We identified 22,658 patients with diabetes who met the inclusion criteria (Fig 1). After excluding individuals who did not have information available on biological sex or SDI or had baseline eosinophil counts exceeding 1,500 cells/μL, the remaining 21,447 patients were included in the primary analysis (Table 1).

**Table 1.** Baseline characteristics of the primary analytic cohort.

| Characteristic | Mean (SD) or N (%) |
| --- | --- |
| *N* | **21,447** |
| Age, years | 60.7 (14.3) |
| Female | 10,274 (47.9%) |
| White | 14,150 (66.0%) |
| Partnered | 10,366 (48.3%) |
| English as primary language | 17,997 (83.9%) |
| Government Insurance | 14,995 (69.9%) |
| Veteran | 2,083 (9.7%) |
| Social Deprivation Index score | 49.1 (32.6) |
| Charlson Comorbidity Index | 9.9 (5.0) |
| Baseline Steroid use | 1,499 (7.0%) |
| BMI<sup>a</sup>, kg/m<sup>2</sup> | 33.0 (7.6) |
| Baseline HbA1c, % | 8.4 (1.8) |
| Baseline Eosinophil count, cells/μL | 186.3 (153.7) |

Categorical variables are presented as n (%), and continuous variables are presented as mean (SD).
The only variable that had missing data was BMI: 11,910 (55.5%) of patients had missing BMI information.
<sup>a</sup> BMI, body mass index.

**Fig 1.**
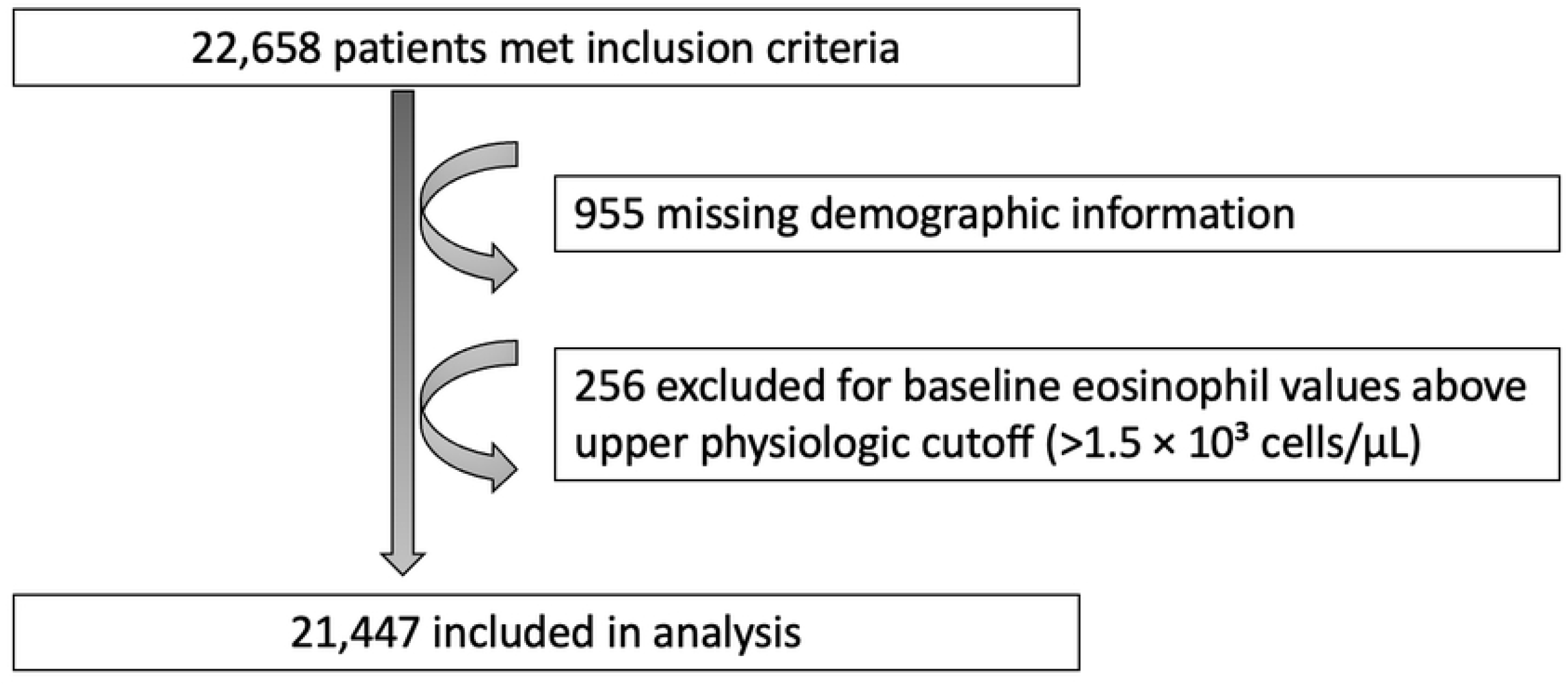
Patient Flow

In this study population, the mean HbA1c decreased from baseline by 0.82% (SD 2.09%) over the 1-year follow-up period. The mean eosinophil count increased by 2.6 cells/μL (SD 157.2 cells/μL) over the same period of time. Mean HbA1c declined progressively across quartiles of eosinophil count change, from a decrease of 0.66% in quartile 1, corresponding to changes in eosinophil counts between -1380 and -61 cells/μL, to a decrease of 1.05% in quartile 4, corresponding to changes in eosinophil counts between 61 and 1330 cells/μL (P < 0.001) (Fig 2).

**Fig 2.**
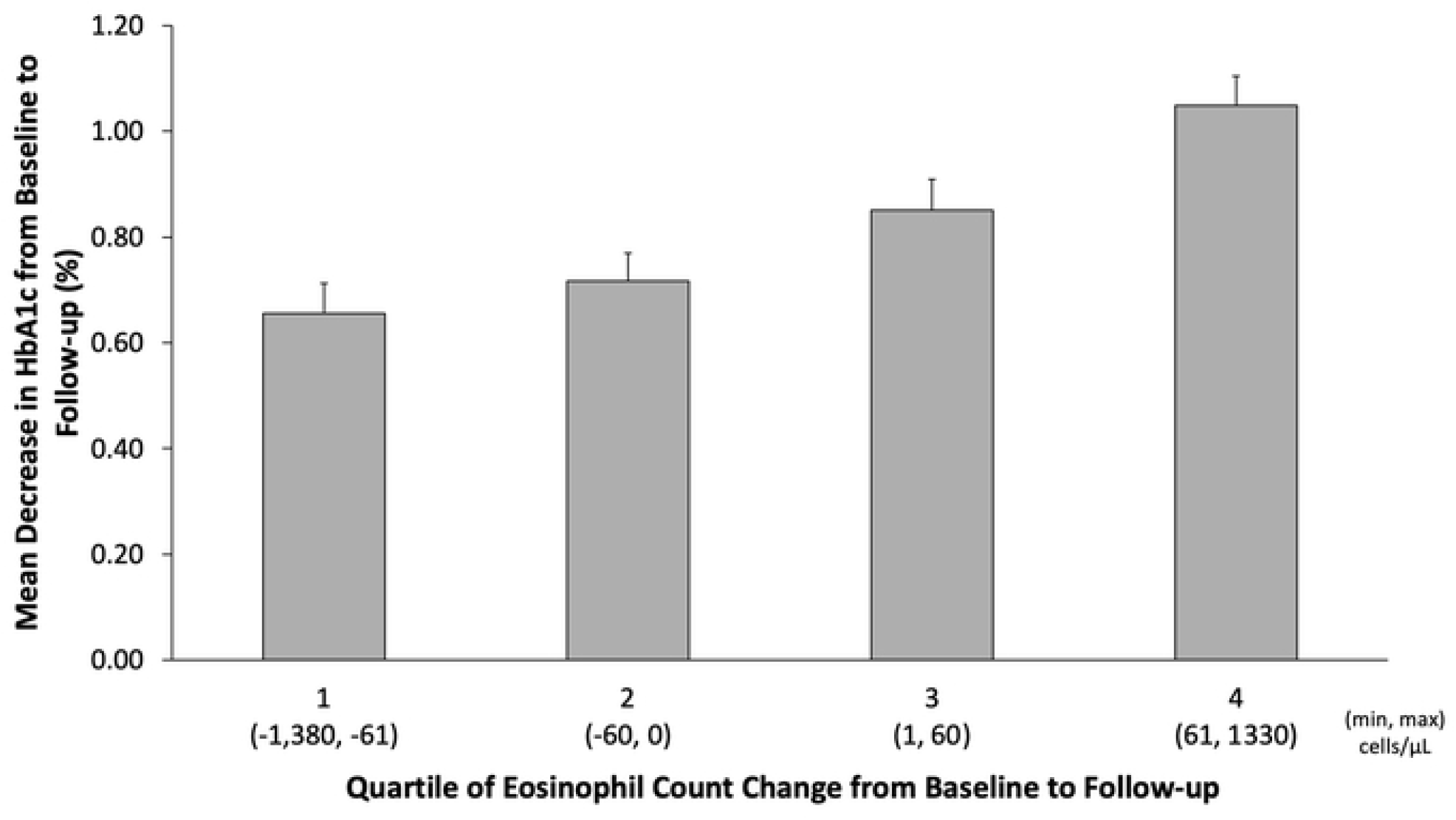
Mean Decrease in HbA1c by Eosinophil Count Change Quartile

In the primary multivariable analysis (Table 2), greater increase in eosinophil count from baseline to follow-up remained associated with a greater decrease in HbA1c (parameter estimate -0.366 per 1,000 cells/μL; 95% CI [-0.526, -0.206]; P < 0.001). Baseline eosinophil count was not significantly associated with a change in HbA1c. Higher baseline HbA1c, older age, White race, and being partnered were associated with a greater decrease in HbA1c; while having government insurance, veteran status, and higher SDI were associated with an increase in HbA1c (Table 2).

**Table 2.** Primary multivariable model results, evaluating change in HbA1c over one year.

| Variable | Estimate | 95% CI | P-value |
| --- | --- | --- | --- |
| Age, per 10 years | -0.154 | -0.174, -0.134 | <0.001 |
| Female | 0.034 | -0.014, 0.081 | 0.170 |
| White | -0.143 | -0.197, -0.088 | <0.001 |
| Partnered | -0.100 | -0.146, -0.054 | <0.001 |
| English as primary language | -0.037 | -0.103, 0.029 | 0.267 |
| Government Insurance | 0.128 | 0.076, 0.180 | <0.001 |
| Veteran | 0.083 | 0.003, 0.163 | 0.042 |
| SDI<sup>b</sup>, per 10 points | 0.018 | 0.010, 0.025 | <0.001 |
| CCI<sup>c</sup> | -0.004 | -0.009, 0.001 | 0.134 |
| BMI<sup>a</sup>, kg/m<sup>2</sup> | -0.010 | -0.015, -0.004 | 0.002 |
| HbA1c, % | -0.764 | -0.777, -0.751 | <0.001 |
| Eosinophil count, per 1,000 cells/μL | -0.041 | -0.207, 0.125 | 0.627 |
| Eosinophil count increase*, per 1,000 cells/μL | -0.366 | -0.526, -0.206 | <0.001 |

<sup>a</sup> BMI, body mass index.
<sup>b</sup> SDI, Social Deprivation Index.
<sup>c</sup> CCI, Charlson Comorbidity Index.
\* over 1 year after study entry.

In age-stratified analyses, greater increase in eosinophil count remained associated with a greater decrease in HbA1c among patients younger than 65 years (parameter estimate -0.487 per 1,000 cells/μL; 95% CI -0.725 to -0.249; P < 0.001) but not among individuals 65 years or older (parameter estimate -0.171 per 1,000 cells/μL; 95% CI -0.364 to 0.022; P = 0.082). In the sensitivity analysis excluding patients who used systemic glucocorticoids either in the year prior to study entry or during follow-up, 19,170 patients remained in the analysis. In this population, greater increase in eosinophil count from baseline to follow-up remained associated with a greater decrease in HbA1c (parameter estimate -0.394 per 1,000 cells/μL; 95% CI -0.567 to -0.221; P < 0.001) whereas baseline eosinophil count was not significantly associated with a change in HbA1c.

Among 13,821 patients with available blood pressure data, mean SBP change did not differ across quartiles of eosinophil count change (P = 0.726). In adjusted analysis, neither baseline eosinophil count nor eosinophil count change was associated with SBP change. Among 13,432 patients with available weight data, patients had progressively less weight loss from quartile 1 (−5.78 pounds) to quartile 4 (−2.66 pounds) of eosinophil count change (P < 0.001). In adjusted analysis, both higher baseline eosinophil count (parameter estimate 3.93 pounds per 1,000 cells/μL; 95% CI 1.64 to 6.22; P < 0.001) and greater increase in eosinophil count (parameter estimate 5.29 pounds per 1,000 cells/μL; 95% CI 2.99 to 7.58; P < 0.001) were associated with greater weight gain / lower weight loss.

## Discussion

In this retrospective cohort of adults with uncontrolled diabetes, greater increases in peripheral eosinophil count were independently associated with greater reductions in HbA1c during follow-up. This association persisted after adjustment for baseline HbA1c and other clinical and sociodemographic characteristics and in a sensitivity analysis excluding patients with documented systemic glucocorticoid exposure. These findings do not establish a causal effect of eosinophils on glycemia. However, they extend prior cross-sectional observations by demonstrating that changes in eosinophil count over time, rather than baseline values, are associated with longitudinal changes in glycemic control.

The mechanisms underlying this association remain uncertain. Experimental studies have shown that adipose-tissue eosinophils secrete IL-4 and IL-13, promoting M2 macrophages that help maintain an anti-inflammatory adipose tissue environment associated with improved insulin sensitivity and glucose homeostasis in mice (5). Human findings have been less consistent. Zhu et al. reported that higher eosinophil percentage was associated with lower prevalence of type 2 diabetes and lower insulin resistance (7), whereas Amini et al. found that higher circulating eosinophil counts were associated with obesity, metabolic syndrome, and higher HbA1c, but not with fasting glucose, type 2 diabetes, or blood pressure (8). Together, these findings suggest that circulating eosinophil counts may not directly reflect the biologic functions of eosinophils within adipose tissue and that their metabolic effects likely depend on tissue location, cellular phenotype, and the surrounding immune environment (11). Differences in study populations, designs, and eosinophil measures may therefore contribute to the apparently conflicting results. Our findings add a distinct longitudinal perspective, suggesting that change in circulating eosinophil count may be more closely associated with glycemic change than a single baseline measurement.

The secondary analyses further underscore the complexity of this relationship. One potential explanation for the observed association between increasing eosinophil count and improving glycemic control is that eosinophil increases reflect reduced endogenous glucocorticoid activity, which would be expected to improve glycemia while also lowering blood pressure and reducing weight gain. However, our findings do not support this hypothesis. Although greater increases in eosinophil count were associated with greater reductions in HbA1c, they were also linked to lower weight loss and had no relationship to changes in systolic blood pressure. Thus, the observed association with HbA1c did not occur alongside parallel improvements across other cardiometabolic parameters and should not be interpreted as evidence that higher eosinophil counts are uniformly metabolically beneficial.

The association between eosinophil-count change and HbA1c reduction appeared stronger among patients younger than 65 years and did not reach statistical significance among older patients. Similar age-related attenuation has been reported for several cardiometabolic biomarkers. For example, associations of the triglyceride-glucose index with incident diabetes and mortality were stronger, or present only, among adults younger than 65 years, while liver fibrosis scores also showed substantially stronger relative associations with mortality in younger than older adults with type 2 diabetes (12-14). These observations suggest that the clinical significance of metabolic biomarkers may vary with age, potentially because older adults have greater heterogeneity in diabetes duration, comorbidity, treatment, and other factors that influence glycemic control. However, because our analyses were stratified and no formal age-by-eosinophil interaction was tested, this finding should be considered exploratory.

Strengths of this study include its large longitudinal cohort, assessment of longitudinal changes in both eosinophil count and HbA1c, adjustment for multiple potential confounders, and sensitivity analysis excluding systemic glucocorticoid users. At the same time, several limitations should be acknowledged. Observational design precludes causal inference, and residual confounding may remain from changes in diabetes treatment, medication adherence, diet, physical activity, smoking, allergic or inflammatory conditions, infection, and other factors that may affect eosinophil counts or HbA1c. Measurements were obtained during routine clinical care rather than at standardized intervals, and circulating eosinophil counts may not reflect eosinophil abundance or function in adipose or other metabolically relevant tissues. Missing BMI values required multiple imputation, and the weight and blood-pressure analyses were restricted to patients with available measurements and may therefore be subject to selection bias. Requiring repeated eosinophil and HbA1c measurements may also have selected patients with greater healthcare utilization. Overall, these findings identify longitudinal eosinophil-count change as an independent correlate of HbA1c change and support prospective studies to determine whether it reflects treatment response, underlying immune-metabolic processes, or another factor associated with improving glycemic control.

## Author Contributions

SB prepared the data, performed the analyses, and drafted the manuscript. AT designed the study and obtained funding. FM assisted with data acquisition. MS assisted with statistical analyses. All authors read and approved the final manuscript for publication.

## Data Availability

Raw patient-level data used in this study cannot be shared openly to protect patient privacy, based on the Mass General Brigham institutional policies. De-identified patient-level data are available under a Data Use Agreement (DUA), in accordance with the Mass General Brigham institutional policies. Data requests can be made to the Mass General Brigham Institutional Review Board at.

